# Integrating Genetic and Transcriptomic Data to Identify Genes Underlying Obesity Risk Loci

**DOI:** 10.1101/2024.06.11.24308730

**Authors:** Hanfei Xu, Shreyash Gupta, Ian Dinsmore, Abbey Kollu, Anne Marie Cawley, Mohammad Y. Anwar, Hung-Hsin Chen, Lauren E. Petty, Sudha Seshadri, Misa Graff, Piper Below, Jennifer A. Brody, Geetha Chittoor, Susan P. Fisher-Hoch, Nancy L. Heard-Costa, Daniel Levy, Honghuang Lin, Ruth JF. Loos, Joseph B. Mccormick, Jerome I. Rotter, Tooraj Mirshahi, Christopher D. Still, Anita Destefano, L. Adrienne Cupples, Karen L Mohlke, Kari E. North, Anne E. Justice, Ching-Ti Liu

## Abstract

Genome-wide association studies (GWAS) have identified numerous body mass index (BMI) loci. However, most underlying mechanisms from risk locus to BMI remain unknown. Leveraging omics data through integrative analyses could provide more comprehensive views of biological pathways on BMI. We analyzed genotype and blood gene expression data in up to 5,619 samples from the Framingham Heart Study (FHS). Using 3,992 single nucleotide polymorphisms (SNPs) at 97 BMI loci and 20,692 transcripts within 1 Mb, we performed separate association analyses of transcript with BMI and SNP with transcript (P_BMI_ and P_SNP_, respectively) and then a correlated meta-analysis between the full summary data sets (P_META_). We identified transcripts that met Bonferroni-corrected significance for each omic, were more significant in the correlated meta-analysis than each omic, and were at least nominally associated with BMI in FHS data. Among 308 significant SNP-transcript-BMI associations, we identified seven genes (*NT5C2*, *GSTM3*, *SNAPC3*, *SPNS1*, *TMEM245*, *YPEL3*, and *ZNF646*) in five association regions. Using an independent sample of blood gene expression data, we validated results for *SNAPC3* and *YPEL3*. We tested for generalization of these associations in hypothalamus, nucleus accumbens, and liver and observed significant (P_META_<0.05 & P_META_<P_SNP_ & P_META_<P_BMI_) results for *YPEL3* in nucleus accumbens and *NT5C2*, *SNAPC3*, *TMEM245*, *YPEL3*, and *ZNF646* in liver. The identified genes help link the genetic variation at obesity risk loci to biological mechanisms and health outcomes, thus translating GWAS findings to function.

## INTRODUCTION

Obesity is an enormous global public health burden.^1^ Since obesity is a major risk factor for numerous health outcomes, including cardiometabolic diseases^2^, the rapid increase in the global obesity burden requires immediate public health action and a better understanding of obesity pathogenicity to prevent it. Decades of research, including genome-wide association studies (GWAS), have demonstrated the fundamental role of genetic susceptibility in obesity risk.^3–13^ Each GWAS-identified locus potentially provides novel biologic insight; yet identifying the functional variants, genes, and underlying pathways at these loci has limited translation for precision medicine.

A major barrier to precision medicine for obesity has been the identification of functional genes underlying GWAS findings. Of the thousands of genomic regions associated with obesity-related traits by GWAS, over 90% are in non-coding, potentially regulatory regions of the genome.^14^ Previous work mapping body mass index (BMI)-related genes implicates the involvement of synaptic function and glutamate receptor signaling, which impinge on key hypothalamic circuits that respond to changes in feeding and fasting and are regulated by key obesity-related molecules such as *BDNF* and *MC4R*^15–18^. These pathways overlap with a proposed mechanism of action of topiramate, a component of one new FDA-approved weight-loss drug^19,20^. However, our understanding of the fundamental mechanisms underlying genetic risk for obesity is limited and controversial even for *FTO*, with the most prominent effects on BMI.^21^

Transcriptomics lie along pathways linking genetic susceptibility to obesity and is emerging as powerful disease biomarkers^22–25^ that may provide targetable “mechanistic bridges” linking GWAS findings with obesity risk. Large-scale characterization and integration of OMICs have been challenging because the comprehensive collection of molecular data has, until very recently, either been unavailable or cost-prohibitive in the context of a single study. However, OMICs scans in the same individuals in which obesity-associated loci discoveries were made are now available^24,26–30^, thereby facilitating comprehensive and efficient integration with genetic data to illuminate the underlying genes and mechanistic pathways of obesity-associated loci. Thus, studies that integrate GWAS with transcriptomics may lead to breakthroughs that reveal the genes contributing to obesity,^31–33^ identify individuals or groups that could benefit from aggressive prevention or treatment, or the repurposing of therapeutics.^34,35^

In this study, we analyzed GWAS data and transcriptomic data generated in whole blood in 5,619 participants from the Framingham Heart Study (FHS) to identify potential causal genes through which known loci operate on obesity phenotypes (BMI). We used a correlated meta-analysis procedure to efficiently screen loci for potential candidate genes that are jointly associated with BMI and SNPs in linkage disequilibrium (LD) with established BMI-associated GWAS SNPs.

## METHODS

### Study sample

We included participants from both the Offspring cohort and the third generation (Gen3) cohort of the FHS. The Offspring cohort of FHS began in 1971 and consisted of children of the Original cohort and spouses of these children.^36,37^ Gen3 cohort comprised children from the offspring families enrolled in 2002.^38^ The time intervals between clinical examinations for Offspring and Gen3 cohorts were approximately 4-6 years.

Since the timing of the blood sample taken for RNA collection was close to the eighth clinical examination (Exam 8) for the Offspring cohort and the second clinical examination (Exam 2) for the Gen3 cohort, our study was restricted to subjects with available blood sample, genotype data, and BMI information in either Exam 8 of the Offspring study or Exam 2 of the Gen3 study. All participants provided written informed consent. The Institutional Review Board of the Boston University Medical Campus approved the study (N=5,169). All participants provided written informed consent. The Institutional Review Board of the Boston University Medical Campus approved the study.

### Data description

FHS participants were genotyped using the Affymetrix GeneChip Human Mapping 500K Array Set and another Affymetrix 50K gene-centric array. The genotype imputation was performed using the Michigan Imputation Server with HRC reference panel release 1.1 April 2016 (HRC r1.1).

Fasting peripheral whole blood samples (2.5 ml) were collected from FHS participants at the eighth clinical examination (Exam 8) of the Offspring cohort and the second clinical examination (Exam 2) of the Gen3 cohort. The details of RNA collection and expression data cleaning have been previously described.^39^ In our study, we used the expression data that have been adjusted using technical covariates and blood count^40,41^.

Height and weight were measured at Exam 8 of the Offspring cohort and Exam 2 of the Gen3 cohort. BMI was then calculated by weight (kg)/height(m)^2^.

### SNP-transcript association and transcript-BMI association

We analyzed 3,992 SNPs that are in LD (r^2^ >0.8) with 97 previously reported BMI variants from GIANT BMI GWAS paper (Locke et al. 2015) and the 1,408 transcripts with a start position within 1 Mb of these variants.

We performed two kinds of association modeling. The first was a SNP-transcript association model, with the transcript as the outcome, and the SNP genotype as the predictor, adjusting for covariates including age at expression data collection, sex, and cohort identifier. We performed this first model for every SNP-transcript pair, using a linear mixed effects model to account for relatedness. The second model assessed the association between transcript and BMI, with expression of the transcript as the outcome, and BMI as the predictor, adjusting for age at expression data collection, sex, cohort identifier, and familial relatedness. We performed the second model for each transcript separately. In this manuscript, we will denote the p-value of the SNP from the first model as P_SNP_ and the p-value of BMI from the second model as P_BMI_.

### Correlated meta-analysis

We used the correlated meta-analysis model of Province and Borecki^42^ to account for the potential dependence between the SNP-transcript and transcript-BMI associations. This correlated meta-analysis model estimated the degree of correlation between SNP-transcript and transcript-BMI associations, and corrected for the inflation of type-I error that would be observed in a traditional meta-analysis (that assumes the two associations are statistically independent). Our model used a tetrachoric correlation, which was less sensitive to contamination from the alternative hypothesis than the Pearson correlation, thus preventing over-correction of the correlation.

In our analysis, for every SNP, we estimated the covariance matrix Σ between two association results 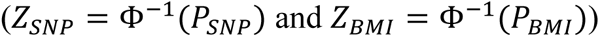 using tetrachoric correlation, and then we calculated 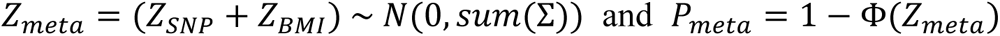 for each SNP-transcript pair.

After performing the correlated meta-analysis, we further screened the results to identify transcripts that met Bonferroni-corrected significance for each omic and were more significant in the correlated meta-analysis than in each omic. Thus, we included five criteria: P_meta_<P_SNP_, P_meta_<P_BMI_, P_SNP_< (0.05/1,408) =3.6×10^-5^, P_BMI_<3.6×10^-5^, and at least nominal association (P<.05) between BMI and SNP in FHS. The first two criteria ensured that both the SNP-transcript and transcript-BMI associations contributed to the meta-analysis. The third and fourth criteria guaranteed the Bonferroni-corrected significance of each association. The last criterion restricted the SNPs to those at least nominally associated with BMI in FHS data.

### Functional interrogation/regulatory annotation

Regulatory variants are more likely to drive correlated signals of gene expression and SNP association than coding variants. To characterize candidate regulatory variants, we used chromatin marks and other epigenomic data that define regulatory elements or link regulatory elements to gene transcription start sites. We focused on data sets for liver, and component cell types, especially preadipocytes, adipocytes, and hepatocytes. We compared them to other tissues because tissue-restricted regulatory elements may be more likely to be relevant and functional. The resources we considered include accessible chromatin based on the assay for transposase-accessible chromatin (ATAC-seq) or DNase hypersensitivity from brain, blood, and liver; histone mark and transcription factor ChIP-seq and chromatin states from ENCODE^43^ used for visual inspection and to assess variant overlap with potential candidate cis regulatory elements (cCREs). Additional resources used for variant annotation as described in Box 1 include GeneCards^44^, OMIM^45^, and GTEx^46^.

### Validation: Cameron County Hispanic Cohort (CCHC)

The CCHC was established on the Texas-Mexico border in 2004.^47^ This randomly ascertained community cohort currently comprises over 5000 people and is approximately 60% female. The mean age of the CCHC participants was 45.2 years and 61% were > age 40. All CCHC individuals were genotyped using the Illumina MEGAEX array.^48^ All genotype data was quality controlled (QC’ed) following standard protocols (i.e. exclude individuals and variants with high levels of missingness, extreme heterozygosity, sex mismatch, variants with low minor allele frequency (MAF <0.01), or those that deviate from Hardy-Weinberg equilibrium (p <10^-6^), then imputed to TOPMed R2 panel.^49^ The study was approved by the Committee for the Protection of Human Subjects (CPHS) at the University of Texas Health Sciences Center at Houston. All participants provided informed consent to be included in CCHC genomic studies.

RNA sequencing of 1,800 CCHC participants was conducted using stored whole blood with sufficient quantity and quality. Sample collection and transcriptome profiling were described in detail previously.^50^ Briefly, pooled libraries were subjected to 150 bp paired-end sequencing according to the protocol (Illumina NovaSeq) at VANTAGE. Six blood cell types were predicted and scaled using the R package DeconCell, a method that quantifies cell types using expression of marker genes.^51^ We performed fastp for quality control following standard protocols,^52^ and the QC-passed reads were aligned to human genome reference (hg38) with STAR.^53,54^ Samples with aligned reads <15M, alignment rate <40%, or assigned reads <15M were excluded.^55^ DESeq2 was performed for library size normalization and gene-specific dispersion estimation.^56^ PEER factor analysis was used to capture the unobserved confounders of transcriptome profiles.^57^ We then implemented a negative binomial model in DESeq2 to identify BMI genes with covariate adjustment for sex, age, 10 PEER factors^56^, and filtered results using default thresholds.

We performed eQTL mapping using the GTEx v8 pipeline.^46^ In brief, we selected 645 unrelated CCHC individuals with both genotyping and RNA-seq data available.^58^ We aligned the RNA-seq reads to the human genome reference using STAR, and then quantified each gene using RNA-SeQC.^53,59^ Read counts were normalized by trimmed mean of M values (TMM), and inverse variance normalization was performed.^60^ We identified eQTLs in cis (within 1 Mb) for each gene using FastQTL with adjustment for sex, RNA-seq batch, 5 genetic principal components (PCs), and 10 PEER factors.

### Generalization on liver tissue: MyCode Bariatric Surgery Cohort

The MyCode™Community Health Initiative (MyCode) study is a healthcare-based population study in central and northeastern Pennsylvania with ∼2 million patients ^61,62^. All participants provided informed consent for the MyCode Study. This study was approved by the Geisinger Institutional Review Board. We leveraged existing transcriptomic profiling in the Geisinger Health System’s (GHS) Bariatric Surgery Program (BSP) study to generalize observed joint associations from whole blood in FHS to liver tissue. All BSP participant data are linked to clinical and demographic data through Mycode’s electronic health records. While the BSP study participants are all obese, there is significant variation across the study participants (**Supplementary Table 1**, N=2,224). Liver wedge biopsies were obtained intraoperatively during Roux-en-Y gastric bypass (RYGB) surgery^63^, as described previously^64,65^. Total RNA was isolated from the excised liver tissue using the RNeasy total RNA isolation kit (Qiagen, Valencia, CA). Standard library prep procedures were followed by sequencing to obtain raw reads for each sample. The sequencing reads were aligned to Ensembl Release 104 reference genotypes and duplicate reads were marked using STAR v2.7.0,^53^ and then quality-controlled and gene expression quantified using and converted to transcripts per million (TPM) using RNASeQC v2.4.2 ^59^. TPM ≥ 0.1 in at least 20% of samples and ≥6 reads in at least 20% of samples. Raw TPM values were inverse normal transformed^60^. PEER factor analysis was used to capture the unobserved confounders of transcriptome profiles.^57^ Association analyses were performed using FastQTL ^66^, adjusting for sex, age, self-identified race/ethnicity, the first three genomic PCs to control for ancestry, and 60 PEER factors.^56^

### Generalization on brain tissue

Analyses of hypothalamus (N=131) and nucleus accumbens (N=198) were conducted on samples from three cohorts: the Framingham Heart Study (FHS), the Religious Orders Study (ROS), and the Rush Memory and Aging Project (MAP). We restricted our analysis to samples with RIN>3 and BRAAK score <= 4. Details of RNA sequencing of hypothalamus and nucleus accumbens and the transcript-BMI association analysis were described previously^67^. For the eQTL analysis, we used FastQTL and adjusted for covariates: 5 first genetic PCs, PEER factors according to the GTEx recommendations (15 PEER factors for hypothalamus and 30 PEER factors for nucleus accumbens), sex, age at death, cohort, and sequencing batch. We further performed meta-analysis using p-values of SNP-transcript and transcript-BMI associations via Fisher’s method^68^, producing a meta-analyzed p-value.

## RESULTS

### Sample characteristics

The characteristics of samples included in the discovery correlated meta-analysis, validation analysis, and generalization analyses are shown in **Supplementary Table 1**. The age distribution was similar for FHS whole blood, CCHC whole blood, and GHS liver analyses, with a mean ranging from 47 to 58, and the brain analyses had relatively older subjects with a mean age of 88. All study samples had a larger proportion of females compared to males. The GHS sample had a relatively higher BMI compared to other study samples. FHS and GHS are dominantly European ancestry, while CCHC was 100% Hispanic/Latino.

### Correlated Meta-analysis

Figure 1 shows the general workflow of our entire study. The models and filtering criteria of each step have been included in the methods section. In the FHS analysis, we found 308 SNP-transcript-BMI associations corresponding to seven unique genes (*NT5C2*, *YPEL3*, *ZNF646*, *SPNS1*, *GSTM3*, *SNAPC3*, and *TMEM245*) potentially involved in transcriptional pathways from SNP to BMI (**Table 1**). 115 variants were involved in the SNP-transcript-BMI associations for *NT5C2*, including the reported BMI variant rs11191560^69,70^. *YPEL3*, *ZNF646* and *SPNS1* were in the same region (16p11.2), and we observed 10, 46 and 91 SNP-transcript-BMI associations for *YPEL3*, *ZNF646* and *SPNS1* respectively, including three reported BMI variants rs4787491 ^28^, rs9925964^69–71^ and rs3888190^28,69,71–74^. At the *TMEM245* locus, we pinpointed the SNP-transcript-BMI association to the reported BMI variant rs6477694^69–71,73–75^. *GSTM3* was located at 1p13.3, with 4 SNP-transcript-BMI associations detected, including previously reported BMI signal rs17024393^28,69,70,76,77^. *SNAPC3*, located at 9p22.3, had 41 SNP-transcript-BMI associations identified. Separate results for suggestive (P_SNP_< (0.05/1,408) =3.6×10^-5^, P_BMI_<3.6×10^-5^) SNP-transcript and transcript-BMI signals are provided in **Supplementary Table 2** and **Supplementary Table 3**.

**Figure 1.**
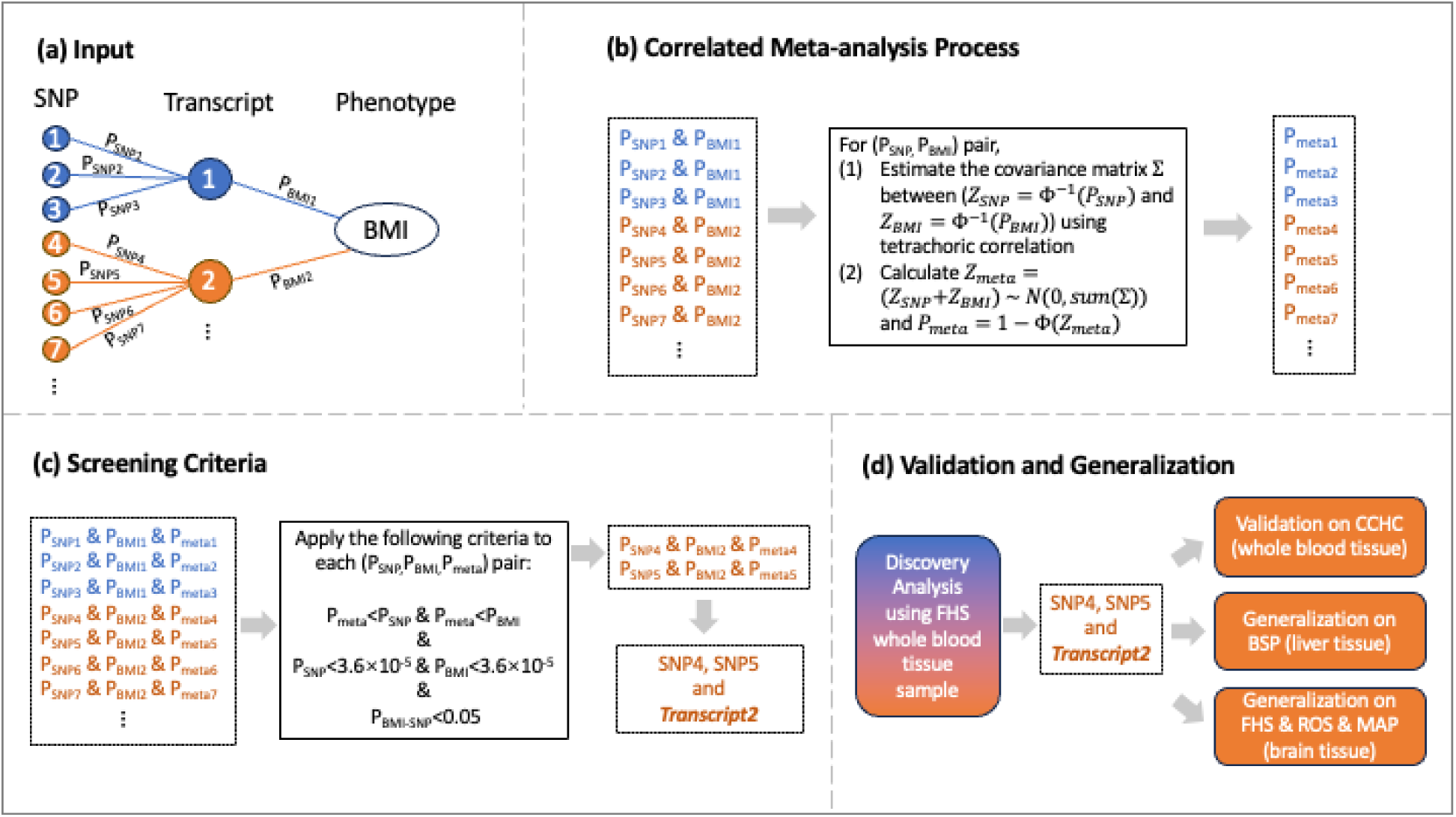
General workflow of the study design. A) Step 1 included single omics associations for SNP to gene expression (P_SNP_) and gene expression to BMI (P_BMI_). B) Step 2 included the correlated meta-analysis to account for the interdependence between P_SNP_ and P_BMI_. C) Identifying all SNP – Gene – BMI combinations that met our filtering criteria, which included correlated meta-analysis results that are more significant than individual omics associations. D) All significant SNP – Gene – BMI combinations were followed by validation in blood, liver, and brain tissues.

**Table 1.** Summary table of significant genes and associations identified in discovery analysis and corresponding validation and generalization results. *Filtering criteria for all validation and generalization analyses: P_META_<0.05 & P_META_<P_SNP_ & P_META_<P_BMI_.

| Gene Symbol | GWAS SNP of locus | Number of SNPs interrogated in FHS | Number of significant SNP-transcript-BMI associations* |  |  |  |  |
| --- | --- | --- | --- | --- | --- | --- | --- |
|  |  |  | Discovery on FHS | Validation on CCHC | Generalization on liver | Generalization on brain – Hypothalamus | Generalization on brain – Nucleus Accumbens |
| *NT5C2* | rs11191560 | 115 | 115 |  | 103 |  |  |
| *GSTM3* | rs17024393 | 13 | 4 |  |  |  |  |
| *SNAPC3* | rs4740619 | 225 | 41 | 37 | 40 |  |  |
| *SPNS1* | rs3888190 | 91 | 91 |  |  |  |  |
| *TMEM245* | rs6477694 | 1 | 1 |  | 1 |  |  |
| *YPEL3* | rs4787491 | 10 | 10 | 10 | 10 |  | 10 |
| *ZNF646* | rs9925964 | 46 | 46 |  | 15 |  |  |

### Validation and Generalization to Other Tissues

We tested for validation of the above seven genes using CCHC blood gene expression data. Among the identified 308 SNP-transcript-BMI associations, 37 SNP-transcript-BMI associations corresponding to *SNAPC3* and 10 SNP-transcript-BMI associations corresponding to *YPEL3* remained significant (P_meta_<0.05 & P_meta_<P_SNP_& P_meta_<P_BMI_) (**Supplementary Table 4**). Regional association plots for each gene show annotation information (Figure 2 and **Supplementary** Figures 1-5). Of note, the top P_META_ SNP for *SNAPC3* and *YPEL3* are within or proximal to putative candidate cis-Regulatory Elements (cCREs) based on ENCODE^43^ regulatory data on blood, brain, and liver tissues (Figure 2).

**Figure 2.**
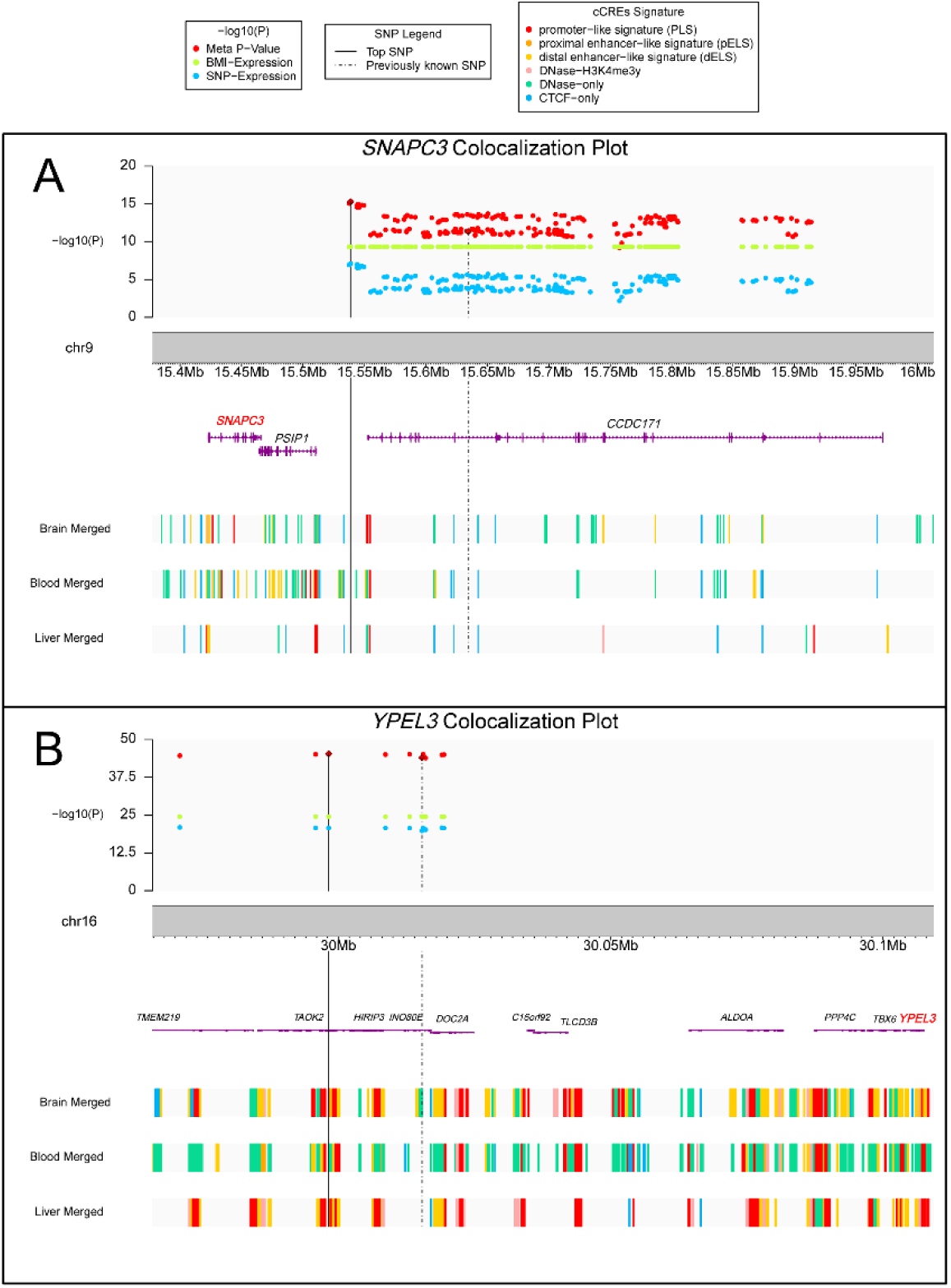
Regional association plot including association results for the discovery sample (Framingham Heart Study) for SNP with gene expression (blue), gene expression with BMI (green), and the correlated meta-analysis for SNP ∼ gene expression ∼ BMI (red). Annotation for potential candidate cis-regulatory elements from ENCODE are included for each reported SNP in the region. A. *SNAPC3*, B. *YPEL3*.

We also tested for generalization using gene expression in brain tissues. Hypothalamus tissue showed no significant SNP-transcript-BMI association. In contrast, the 10 SNP-transcript-BMI associations corresponding to *YPEL3* were significant in the generalization analysis on nucleus accumbens. Additionally, we were able to generalize signals in liver tissue for *NT5C2*, *SNAPC3*, *TMEM245*, *YPEL3*, and *ZNF646*, including 103, 40, 1, 10, and 15 SNP-transcript-BMI associations, respectively (**Supplementary Table 4)**. While the direction of effect was consistent for both brain tissues, even for non-significant associations (**Table 2**, Figure 3, and **Supplementary Table 4**). The direction of effect was not always consistent across tissue types; however, consistency of direction of effect across various tissues may not be expected. Further work may be needed to clarify expectations of directional consistency across tissues with respect to BMI ∼ Gene and SNP ∼ associations.

**Figure 3.**
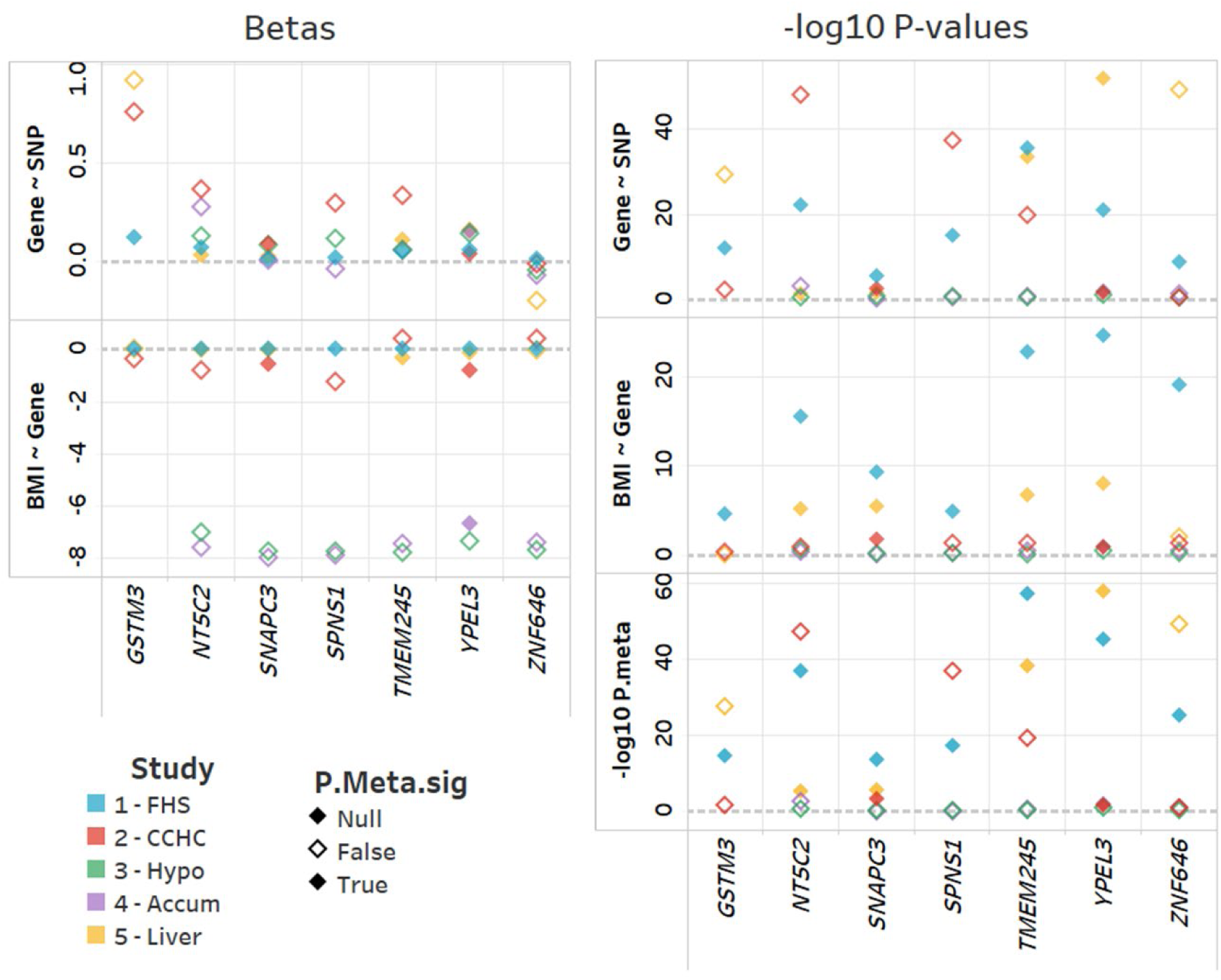
Summary of validation and generalization for most significant SNP in discovery corelated meta-analysis. Results are provided for discovery sample (FHS, blue), validation in blood (CCHC, red), and generalization to hypothalamus (Hypo, green), nucleus accumbens (Accum, purple), and liver (yellow) tissues. We provide individual effect estimates and P-values for each ‘omic and meta-analysis. Filled diamonds indicate significant associations in the meta-analysis (Note: FHS is noted as NULL, as all are significant).

**Table 2.** Summary results for most significant SNP - expression - BMI combination identified in Discovery (FHS) sample. EA – effect allele, OA – other allele.

| Gene | CHR | POS (b38) | dbSNP ID | EA | OA | GWAS tag SNP | GWAS tag dbSNP ID | Study | P<sub>BMI</sub> | N<sub>BMI</sub> | BETA<sub>BMI</sub> | P<sub>SNP</sub> | N<sub>SNP</sub> | BETA<sub>SNP</sub> | P<sub>META</sub> |
| --- | --- | --- | --- | --- | --- | --- | --- | --- | --- | --- | --- | --- | --- | --- | --- |
| *GSTM3* | chr1 | 109612066 | rs17024393 | C | T | Yes | rs17024393 | FHS - Blood | 2.67E-05 | 5,619 | 0.003 | 9.30E-13 | 5,257 | 0.123 | **3.89E-15** |
|  |  |  |  |  |  |  |  | CCHC - Blood | 4.37E-01 | 934 | -0.366 | 5.56E-03 | 769 | 0.757 | 1.70E-02 |
|  |  |  |  |  |  |  |  | Hypothalamus | 9.48E-01 | 131 | -7.858 | - | - | - | - |
|  |  |  |  |  |  |  |  | Nucleus Accumbens | 1.61E-01 | 198 | -7.033 | - | - | - | - |
|  |  |  |  |  |  |  |  | Liver | 7.06E-01 | 2,225 | 0.007 | 6.56E-30 |  | 0.914 | 3.17E-28 |
| *NT5C2* | chr10 | 103154183 | rs74233809 | C | T | No | rs11191560 | FHS - Blood | 3.48E-16 | 5,619 | -0.004 | 5.82E-23 | 5,257 | 0.072 | **1.16E-37** |
|  |  |  |  |  |  |  |  | CCHC - Blood | 1.07E-01 | 934 | -0.803 | 7.98E-49 | 769 | 0.363 | 9.76E-48 |
|  |  |  |  |  |  |  |  | Hypothalamus | 2.02E-01 | 131 | -7.037 | 4.30E-01 | 108 | 0.128 | 2.99E-01 |
|  |  |  |  |  |  |  |  | Nucleus Accumbens | 3.66E-01 | 198 | -7.612 | 7.69E-04 | 170 | 0.276 | 2.59E-03 |
|  |  |  |  |  |  |  |  | Liver | 6.40E-06 | 2,225 | -0.044 | 4.87E-02 | 2,225 | 0.035 | **4.98E-06** |
| *SNAPC3* | chr9 | 15786904 | rs10962158 | A | T | No | rs4740619 | FHS - Blood | 4.71E-10 | 5,619 | 0.002 | 2.97E-06 | 5,257 | 0.015 | **3.66E-14** |
|  |  |  |  |  |  |  |  | CCHC - Blood | 1.63E-02 | 934 | -0.559 | 2.96E-03 | 769 | 0.091 | **5.28E-04** |
|  |  |  |  |  |  |  |  | Hypothalamus | 6.41E-01 | 131 | -7.751 | 2.88E-01 | 108 | 0.083 | 4.97E-01 |
|  |  |  |  |  |  |  |  | Nucleus Accumbens | 7.70E-01 | 198 | -7.979 | 8.19E-01 | 170 | 0.011 | 9.21E-01 |
|  |  |  |  |  |  |  |  | Liver | 4.03E-06 | 2,225 | -0.033 | 3.79E-02 | 2,225 | 0.025 | **2.55E-06** |
| *SPNS1* | chr16 | 28814099 | rs11860513 | T | C | No | rs3888190 | FHS - Blood | 1.33E-05 | 5,619 | -0.001 | 1.03E-15 | 5,257 | 0.019 | **4.59E-18** |
|  |  |  |  |  |  |  |  | CCHC - Blood | 4.12E-02 | 934 | -1.266 | 4.98E-38 | 769 | 0.293 | 1.85E-37 |
|  |  |  |  |  |  |  |  | Hypothalamus | 6.62E-01 | 131 | -7.764 | 2.38E-01 | 108 | 0.119 | 4.49E-01 |
|  |  |  |  |  |  |  |  | Nucleus Accumbens | 6.24E-01 | 198 | -7.902 | 6.37E-01 | 170 | -0.038 | 7.64E-01 |
|  |  |  |  |  |  |  |  | Liver | 6.17E-01 | 2,225 | 0.006 | - | - | - | - |
| *TMEM245* | chr9 | 109170062 | rs6477694 | T | C | Yes | rs6477694 | FHS - Blood | 2.39E-23 | 5,619 | 0.005 | 2.95E-36 | 5,257 | 0.055 | **1.09E-57** |
|  |  |  |  |  |  |  |  | CCHC - Blood | 3.91E-02 | 934 | 0.396 | 2.17E-20 | 769 | 0.337 | 4.20E-20 |
|  |  |  |  |  |  |  |  | Hypothalamus | 7.22E-01 | 131 | -7.796 | 5.13E-01 | 108 | 0.059 | 7.38E-01 |
|  |  |  |  |  |  |  |  | Nucleus Accumbens | 2.88E-01 | 198 | -7.455 | 1.89E-01 | 170 | 0.062 | 2.13E-01 |
|  |  |  |  |  |  |  |  | Liver | **1.72E-07** | 2,225 | -0.341 | 3.78E-34 | 2,225 | 0.113 | **6.08E-39** |
| *YPEL3* | chr16 | 29986879 | rs4077410 | G | A | No | rs4787491 | FHS - Blood | 3.19E-25 | 5,619 | 0.007 | 1.68E-21 | 5,257 | 0.057 | **5.98E-46** |
|  |  |  |  |  |  |  |  | CCHC - Blood | 1.07E-01 | 934 | -0.803 | 3.21E-02 | 769 | 0.037 | **2.30E-02** |
|  |  |  |  |  |  |  |  | Hypothalamus | 3.22E-01 | 131 | -7.365 | 1.07E-01 | 108 | 0.144 | 1.50E-01 |
|  |  |  |  |  |  |  |  | Nucleus Accumbens | 1.04E-01 | 198 | -6.692 | 1.23E-02 | 170 | 0.156 | **9.79E-03** |
|  |  |  |  |  |  |  |  | Liver | **1.09E-08** | 2,225 | -0.127 | 1.39E-52 | 2,225 | 0.163 | **2.10E-58** |
| *ZNF646* | chr16 | 31077026 | rs749671 | A | G | No | rs9925964 | FHS - Blood | 9.68E-20 | 5,619 | -0.003 | 1.89E-09 | 5,257 | 0.017 | **7.33E-26** |
|  |  |  |  |  |  |  |  | CCHC - Blood | 3.91E-02 | 934 | 0.396 | 6.16E-01 | 769 | -0.010 | 1.14E-01 |
|  |  |  |  |  |  |  |  | Hypothalamus | 5.83E-01 | 131 | -7.708 | 4.44E-01 | 108 | -0.042 | 6.09E-01 |
|  |  |  |  |  |  |  |  | Nucleus Accumbens | 2.72E-01 | 198 | -7.414 | 7.26E-02 | 170 | -0.067 | 9.71E-02 |
|  |  |  |  |  |  |  |  | Liver | 9.25E-03 | 2,225 | -0.020 | 7.32E-50 | 2,225 | -0.195 | 8.05E-50 |

### Biological interrogation

Previous studies of gene function and bioinformatics characteristics (see Methods) of the significant genes highlight nearby signatures of gene regulation (**Box 1**, Figure 2 and **Supplementary** Figures 1-5). Additionally, our literature search provides further details on potential roles for identified genes for obesity (**Box 1**). For example, *NT5C2* deletion was found to be protective in mice fed a high fat diet (HFD) ^78^. Other model organism studies have shown alterations in *YPEL3* results in altered obesity phenotypes. For example, *YPEL3* knockdown in *Drosophila melanogaster* resulted in significant changes in body fat percentage^79^. Additionally, previous studies integrating GWAS and eQTL data have shown *YPEL3* and *NT5C2* were both have pleiotropic effects leading to schizophrenia and cardiometabolic disease, schizophrenia and BMI for *YPEL3*, and schizophrenia, BMI, and coronary artery disease for *NT5C2*^80^. Box 1 shows strong support for *YPEL3* and *NT5C2* as likely candidate genes underlying the association with BMI in these two regions. However, existing knowledge that may offer a role for the other genes in the pathway to BMI is sparse.

## DISCUSSION

This study incorporated a correlated meta-analysis method to perform integrative analysis using genotype, gene expression, and phenotype (BMI) data. From the discovery analysis using the FHS whole blood data, we identified seven genes (*NT5C2*, *YPEL3*, *ZNF646*, *SPNS1*, *GSTM3*, *SNAPC3*, and *TMEM245*) that potentially lie along the pathway linking genetic variation to elevated BMI. Among those seven genes, *YPEL3* and *SNAPC3* associations were validated in whole blood in the CCHC study. In the analyses of tissues other than blood, *NT5C2*, *SNAPC3*, *TMEM245*, *YPEL3*, and *ZNF646* associations generalized in the liver tissue, and *YPEL3* in the nucleus accumbens.

*YPEL3* is located at 16p11.2, a gene dense region well-known for a microdeletion associated with neurocognitive developmental delay and predisposition to obesity^81–83^. Literature has reported that this region’s deletion event is related to a highly-penetrant form of obesity^84,85^, and is age- and gender-dependent^86,87^. Within this region, *SH2B1* has received much attention as the likely causal gene underlying the mosaic effects of the 16p11.2 deletion and is thought to regulate body weight and glucose metabolism^88,89^; and as a result, *YPEL3* has rarely been considered in previous studies. One of the previous studies that considered *YPEL3*^80^ identified it as a pleiotropic gene jointly influencing BMI and risk of schizophrenia. In contrast, another study^90^ asserted that the association between *YPEL3* and schizophrenia is due to its correlation with expression of *INO80E*, another possible candidate gene for BMI and risk of schizophrenia in the 16p11.2 region. Despite the controversial findings of *YPEL3* in the literature, several pieces of evidence support a role of *YPEL3* in BMI. First, the gene is highly expressed in whole blood and brain, similar to well-known BMI-related genes (**Box 1**). Also, *YPEL3* was the sole candidate gene in this region identified by the current analysis. Further, the blood expression results were validated in an independent study of Hispanic participants, and the results generalized to both brain and liver tissues. Combined, this evidence suggests that more attention is warranted on this gene in the future.

*NT5C2* is located at 10q24.32, which has been reported as a highlight locus of autism spectrum disorder, brain arterial diameters, and schizophrenia^91–93^. A previous study in zebrafish found *NT5C2* as a potential causal gene in this region for blood pressure^94^. Notably, variation in this gene is also associated with lower visceral and subcutaneous fat^95^, obesity, and the concurrence of obesity and depression^96^ (**Box 1**). Further, animal studies of *NT5C2* knock-outs show changes in body weight gain, insulin resistance on high-fat diet, and white adipose tissue mass^78,97^. Kumar et al. found that rs11191548 decreased miRNA binding efficiency, which may explain the functional role of *NT5C2* influencing BMI^98^. Yet, our significant findings linking SNP variation to *NT5C2* gene expression with BMI in liver tissue is novel and a role for this gene in other tissues warrants further exploration.

While support for other genes identified herein is limited in the literature, *SNAPC3*, which validated in CCHC, and *TMEM245*, which generalized to liver tissue, have connections to obesity-related traits. For example, similar to both genes mentioned above, *SNAPC3* variants have also been associated with schizophrenia^99^. Also, DNA methylation in *SNAPC3* has been reported to mediate the association between breastfeeding and early-life growth trajectories^100^. The expression level of *TMEM245* has been associated with atrial fibrillation^101^, and schizophrenia-associated variants have been reported within this gene^102^.

In recent years, there has been growing interest in developing integrative approaches that utilize various OMICs data to uncover underlying biological mechanisms^103–106^. When individual-level data is available, combining multiple OMICs datasets to perform further analysis is preferred^107,108^. Yet, few integrative studies using summary-level data exist^80,109^, limiting cross study analyses. Thus, among all the integrative OMICs analyses, the correlation between OMICs is often ignored^109,110^. In our study, we leveraged the correlated meta-analysis framework proposed^42^, which is a robust approach to integrate “suspected” correlated SNP-transcript association and transcript-BMI association. This approach is useful for performing statistical integration and has been incorporated into many colocalization and polygenic pleiotropy detection methods^111,112^. By performing correlated meta-analysis using summary level data, we ensured the correlation between summary statistics of OMICs scans were considered.

There are some previous integrative studies on obesity-related phenotypes. Smemo et al^21^. found that obesity-associated variants within FTO were functionally connected with *IRX3 and IRX5* expression. Voisin et al.^113^ and Tang et al.^114^ evaluated the association and the interaction between obesity-associated SNPs and DNA methylation changes. Kogelman et al.^107^ detected co-expression patterns among eQTLs, integrated with protein data, and detected several obesity candidate genes, such as *ENPP1*, *CTSL*, and *ABHD12B*. More recently, integrative analyses on multiple obesity and neuro-related phenotypes provided further gene lists that potentially affected relevant phenotypes jointly^80,90^. Also, a recent study colocalized splice junction quantitative trait loci (sQTLs) measured in subcutaneous adipose tissue with 24 BMI GWAS loci, including with *YPEL3*^115^, and another study has reported 162 BMI signals with a colocalized adipose eQTL.^116^

Compared to other integrative studies, our study has several strengths. To our knowledge, our study is the first one that takes the correlation between OMICs scans into the integrative analysis of BMI. We not only have a discovery study using whole blood samples from European ancestry, but validate these joint associations in an independent study of Hispanic participants, and generalize our findings to other tissues, including liver nucleus accumbens. Yet, our study has some limitations. First, the traditional meta-analysis instead of the correlated meta-analysis was used in the validation and generalization analyses due to data sparsity. Also, we only included two types of OMICs data in our analyses, genetics and gene expression data. However, these analyses gave us a comprehensive view of how our findings can be interpreted across ancestry and tissue type. And, our work offers a framework for future investigations incorporating additional of OMICs data, such as DNA methylation or protein data, as well as additional tissues, that can also be adopted for other traits of interest.

## CONCLUSION

Our study aimed to narrow in on causal genes that underly known obesity susceptibility loci. Specifically, we were interested in genetic variation that may be operating on variation in BMI through alterations in gene expression. Our integrative, multi-omics approach identified seven candidate genes within five genomic regions for BMI. Among these seven, we find the strongest support for *YPEL3*, *NT5C2*, and *SNAPC3*, through validation across ancestries, generalization across BMI-relevant tissues, and/or existing literature with a connection to BMI-related traits or gene functions. This deep dive into the etiology of obesity risk loci gets us one step forward to connecting genetic variation to biological mechanisms and health outcomes, and thus translating GWAS findings to function so that obesity precision treatment and prevention can begin.

## Data Availability

All data produced in the present study are available upon reasonable request to the authors.

## AUTHOR CONTRIBUTIONS

Participate in Project Concept and Design: LAC, KEN, AEJ, CTL. Participate in Parent Study Concept and Design: JBM, JIR, LAC, KEN. Participate in Phenotype Data Acquisition and/or QC: HX, ID, SPFH, RJFL, NHC, JBM, TM, CDS, LAC, AD, KEN, AEJ, CTL. Participate in Genotype or gene expression Data Acquisition and/or QC: HX, ID, MYA, HHC, LEP, MG, GC, HL, NHC, SPFH, AD, RJFL, JBM, TM, CDS, LAC, KEN, AEJ, CTL. Participate in Data Analysis and Interpretation: HX, SG, ID, MYA, RJFL, KLM, KEN, AEJ, CTL. Drafted the manuscript and revised according to co-author suggestions: HX, SG, SPFH, KEN, AEJ, CTL. All authors critically reviewed the manuscript, suggested revisions as needed, and approved the final version.

## ACKNOWLEDGEMENTS

Individual Acknowledgements:

SG, ID, GC, CTL, KEN, AEJ, CDS, TM, KLM were funded in part by NIH R01 DK122503. MYA was funded by NIH NIDDK 3R01DK122503 – 02W1. MG was funded in part by NIH R01HL163262. JIR was funded in part by National Center for Minority Health Disparity (NCMHD), MD000170P20, McCormick. KEN was also funded in part by NIH R01HL142302, R01HL151152, R01 R01HD057194, R01HG010297, and R01HL143885.

**Study Acknowledgements:**

<u>The Framingham Heart Study (FHS)</u>: The FHS is funded by National Institutes of Health contract N01-HC-25195. The laboratory work for this investigation was funded by the Division of Intramural Research, National Heart, Lung, and Blood Institute, National Institutes of Health, Bethesda, MD. The analytical component of this project was funded by the Division of Intramural Research, National Heart, Lung, and Blood Institute, and the Center for Information Technology, National Institutes of Health, Bethesda, MD. The visualization tools and data resources for this project were funded by the National Center for Biotechnology Information, National Institutes of Health, Bethesda, MD.

The FHS acknowledges the support of contracts NO1-HC-25195, HHSN268201500001I and 75N92019D00031 from the National Heart, Lung and Blood Institute and grant supplement R01 HL092577-06S1 for this research. We also acknowledge the dedication of the FHS study participants without whom this research would not be possible. This manuscript does not necessarily reflect the opinions or views of the NHLBI, NIH or DHHS.

<u>Cameron County Hispanic Community (CCHC) Cohort</u>: We thank our cohort team including the CRU, data and lab staff of the CCHC team. We thank Valley Baptist Medical Center, Brownsville, for providing the space for our Clinical Research Unit. CCHC study and team members were funded by: National Institutes of Health, National Center for Minority Health Disparity, MD000170P20, UL1 TR000371, National Heart, Lung, and Blood Institute, 1R01HL142302, 2R01HL142302, National Center for Advancing Translational Sciences, (NCATS), Clinical and Translational Science Awards (CTSA),UL1 9/30/06-6/30/11, UL1TR0003716/27/12-/31/17, UL1TR0031677/1/19-6/30/2

<u>MyCode Bariatric Surgery Cohort</u>: We thank all the participants of the MyCode Community Health Initiative Study (MyCode) and the MyCode Research Team. We thank the members of the Geisinger-Regeneron DiscovEHR Collaboration who have been critical in the generation of the genetic and transcriptomic data used in this study. This study was funded in part by NIH HL142302, NIH 2R01HL142302, National Center for Advancing Translational Sciences, UL1 9/30/06-6/30/11, UL1TR0003716/27/12-/31/17, UL1TR0031677/1/19-(NCATS), Clinical and Translational Science Awards (CTSA).

## CONFLICTS OF INTEREST

None to report.

BOX

## Box 1.

Seven genes identified as significant in the discovery analysis

***NT5C2. 5’-nucleotidase, cytosolic II*** is a protein-coding gene that may maintain internal composition of nucleotides. It hydrolyzes IMP (inosine monophosphate) and other purines (Genecards). *Chr10:104,845,940-104,953,056* (GRCh37/hg19 by Ensembl^117^). No known monogenic conditions reported in OMIM (OMIM). *NT5C2* is ubiquitously expressed (GTEx) with the highest expression observed in the thyroid and esophagus. Mouse knockout models demonstrate reduced body weight gain, insulin resistance on high-fat diet, and white adipose tissue mass ^78,97^**. In-vitro studies in human skeletal muscle tissue show a** suppression of 5’-Nucleotidase enzymes that promote AMP-activated protein kinase (AMPK) phosphorylation and metabolism^118^, which may suggest metabolic flexibility in an obese state. Genetic variations in *NT5C2* have been associated with lower visceral and subcutaneous fat^95^, obesity, and the concurrence of obesity and depression^96^. Two GWAS in East Asian populations identified *NT5C2,* rs113278154, as “associated with metabolically unhealthy phenotypes among normal weight individuals”^119,120^. Rs11191548 of *NT5C2*, which is in complete LD (All pop: R∧2=.9815) with the GWAS index SNP in the region, rs11191560, was studied to determine if miRNAs in the region disrupted binding to their target gene in an allele specific manner^98^. The study found that rs11191548 altered luciferase activity and decreased miRNA binding efficiency, and thus could explain how *NT5C2* may be one of the functional genes influencing BMI.

***YPEL3. Yippee-Like 3*** is a protein-coding gene that is involved in the proliferation and apoptosis in myeloid precursor cells (Genecards). It is required for central and peripheral glial cell development, and mutation of *YPEL3* causes neuropathy^121^. *chr16:30,103,635-30,108,236* (GRCh37/hg19 by Ensembl^117^). No known monogenic conditions reported in OMIM (OMIM). *YPEL3* is ubiquitously expressed (GTEx), with the highest expression observed in the whole blood and brain. Mouse knockout has a small body size and has neuronal irregularities, according to the International Mouse Phenotyping Consortium (IMPC). Liu and colleagues in 2020 identified *YPEL3* as a pleiotropic gene jointly influencing BMI and risk of schizophrenia, further supporting a neuronal correlation of this gene for obesity^80^. *YPEL3* knockdown in *Drosophila melanogaster* resulted in significant changes in body fat percentage^79^.

***ZNF646. Zinc Finger Protein 646*** is a protein-coding gene predicted to enable DNA-binding transcription factor activity, RNA polymerase II-specific, and RNA polymerase II cis-regulatory region sequence-specific DNA binding activity. *chr16:31,085,743-31,095,517* (GRCh37/hg19 by [Ensembl](https://grch37.ensembl.org/Homo_sapiens/Gene/Summary?g=ENSG00000167395)). *ZNF646* is ubiquitously expressed (GTEx) with the highest expression observed in the testis. No known monogenic conditions reported in OMIM (OMIM).

Expression of *ZNF646* has been associated with Parkinson’s disease in two different studies^122,123^.

***TMEM245. Transmembrane protein 245*** is a protein-coding gene with no known function. *chr9:111,777,432-111,882,225* (GRCh37/hg19 by Ensembl^117^). *TMEM245* is ubiquitously expressed (GTEx) with the highest expression in the thyroid, ovaries, and adrenal glands. Zhang and colleagues reported an association between *TMEM245* gene expression levels and atrial fibrillation^101^. Variants in this gene have been associated with schizophrenia^102^, age of menarche^124^, body height^77^, and cognitive abilities^125^.

***SPNS1. Lysosomal H(+)-carbohydrate transporter*** is a protein-coding gene that functions in lysosomal recycling at a late stage of autophagy (Genecards). *SPNS1* also functions as a sphingolipid transporter and may be involved in necrotic or autophagic cell death (Genecards). *Chr16:28,985,542-28,995,869* (GRCh37/hg19 by Ensembl^117^). No known monogenic conditions reported in OMIM (OMIM). *SPNS1* is ubiquitously expressed, with the strongest gene expression in arteries and the uterus. Differential expression of this gene has been associated with BMI^126^ . This region has multiple coordinately regulated genes based on eQTL. In a recent in-silico study, *SPNS1* was differentially expressed in persons with T2D and obesity^127^. Polymorphisms in *SPNS1* have been associated with BMI^128^, asthma^128^, allergic disease^129^, and ADHD^130^ in GWAS.

***GSTM3.* Glutathione S-Transferase Mu 3** is a protein coding gene, an enzyme that belongs to the mu class and functions in the detoxification of electrophilic compounds, including carcinogens, therapeutic drugs, environmental toxins, and products of oxidative stress, by conjugation with glutathione. The genes encoding the mu class of enzymes are organized in a gene cluster on chromosome 1p13.3 and are known to be highly polymorphic. *GSTM3* may be involved in the uptake and detoxification of harmful compounds in the body at the testis and blood-brain barrier (Genecards). *chr1:110,276,554-110,284,384* (GRCh37/hg19 by Ensembl^117^). No known monogenic conditions reported in OMIM (OMIM). *GSTM3* is ubiquitously expressed with the strongest expression in the testis and ovaries. *GSTM3* has been associated with hyperinsulinemia, T2D^131^, and hypertension^132^. Recent studies have reported an increase of *GSTM3* in the omental fat of polycystic ovary syndrome (PCOS)^133^.

***SNAPC3. Small nuclear RNA activating complex polypeptide 3 is*** part of the SNAPc complex required for the transcription of both RNA polymerase II and III small-nuclear RNA genes (Genecards). *SNAPC3* binds to the proximal sequence element (PSE), a non-TATA-box basal promoter element common to these 2 types of genes (GeneCard). *Chr9:15,422,702-15,465,951* (GRCh37/hg19 by Ensembl^117^). *SNAPC3* is ubiquitously expressed with the strongest expression in the testis and the cerebellum. Variants in this gene have been associated with schizophrenia^99^. DNA methylation in *SNAPC3* mediates the association between breastfeeding and early-life growth trajectories^100^.

## Notes

### Competing Interest Statement

The authors have declared no competing interest.

### Author Declarations

The Institutional Review Boards of the Boston University Medical Campus and the Geisinger approved the study. The study was approved by the Committee for the Protection of Human Subjects (CPHS) at the University of Texas Health Sciences Center at Houston.

