## Supplemental Figures for "Integrating Genetic and Transcriptomic Data to Identify Genes Underlying Obesity Risk Loci"

**Supplementary Figure 1.** Regional association plot for the *NT5C2* gene including association results for the discovery sample (Framingham Heart Study) for each tested SNP with gene expression (blue), gene expression with BMI (green), and the correlated meta-analysis for SNP ~ gene expression ~ BMI (red). Annotation for potential ccREs from ENCODE are included for the region.

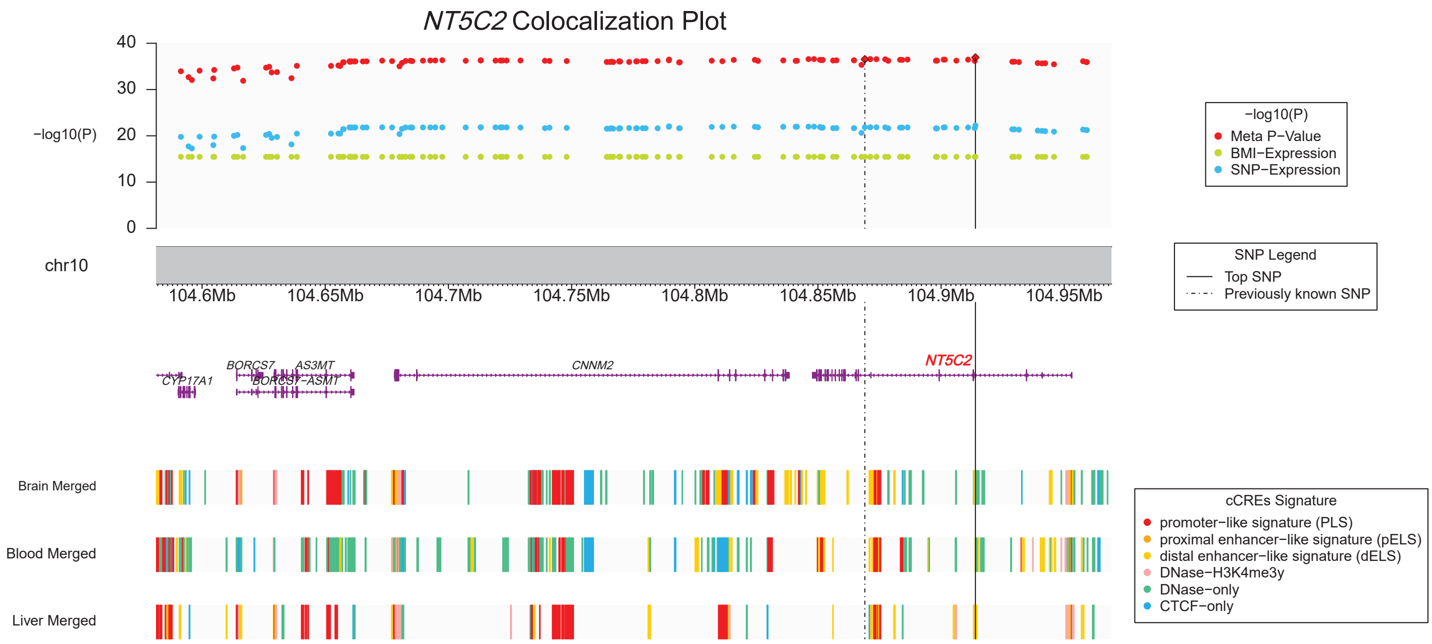

**Supplementary Figure 2.** Regional association plot for the *GSTM3* gene including association results for the discovery sample (Framingham Heart Study) for each tested SNP with gene expression (blue), gene expression with BMI (green), and the correlated meta-analysis for SNP ~ gene expression ~ BMI (red). Annotation for potential ccREs from ENCODE are included for the region.

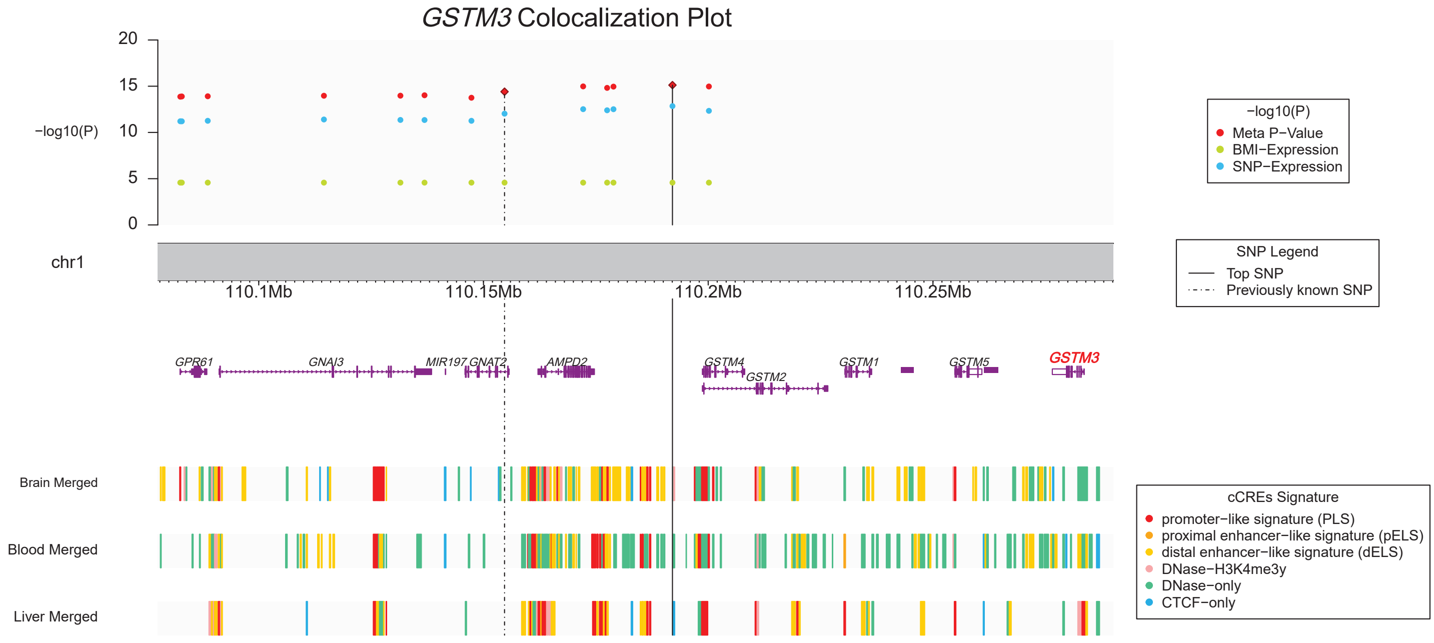

**Supplementary Figure 3.** Regional association plot for the *SPNS1* gene including association results for the discovery sample (Framingham Heart Study) for each tested SNP with gene expression (blue), gene expression with BMI (green), and the correlated meta-analysis for SNP ~ gene expression ~ BMI (red). Annotation for potential ccREs from ENCODE are included for the region.

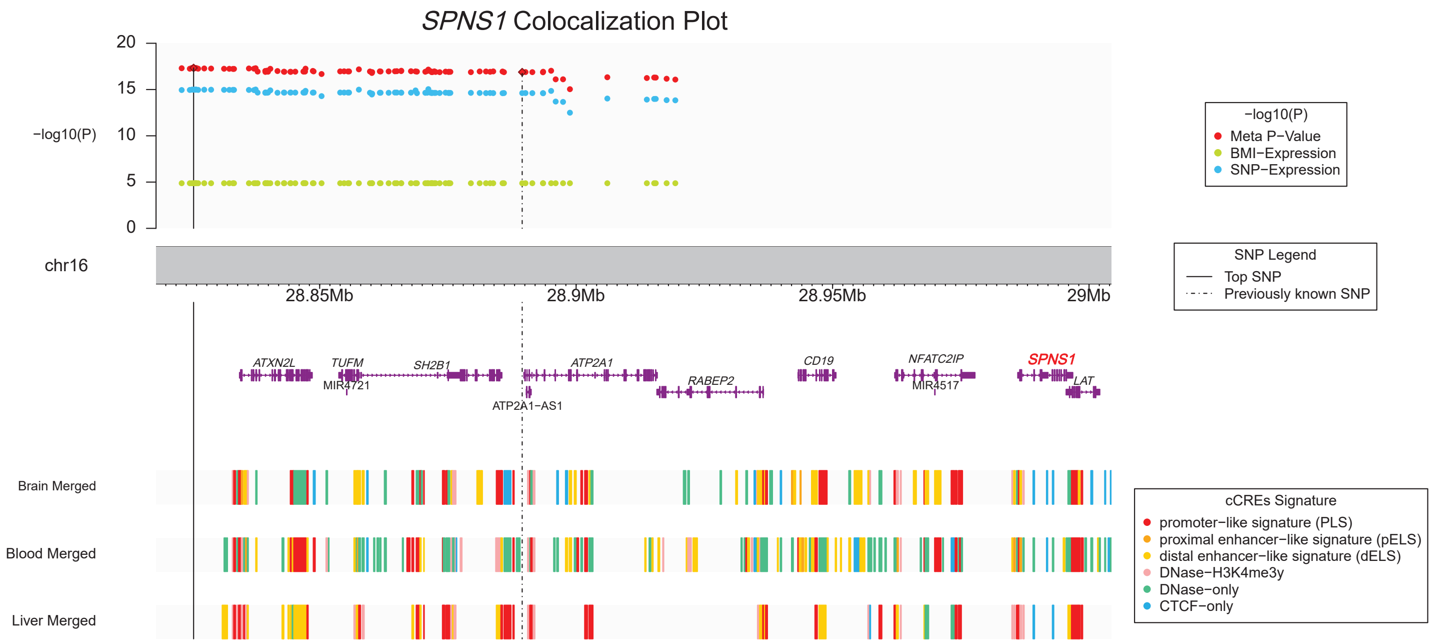

**Supplementary Figure 4.** Regional association plot for the *TMEM245* gene including association results for the discovery sample (Framingham Heart Study) for each tested SNP with gene expression (blue), gene expression with BMI (green), and the correlated meta-analysis for SNP ~ gene expression ~ BMI (red). Annotation for potential ccREs from ENCODE are included for the region.

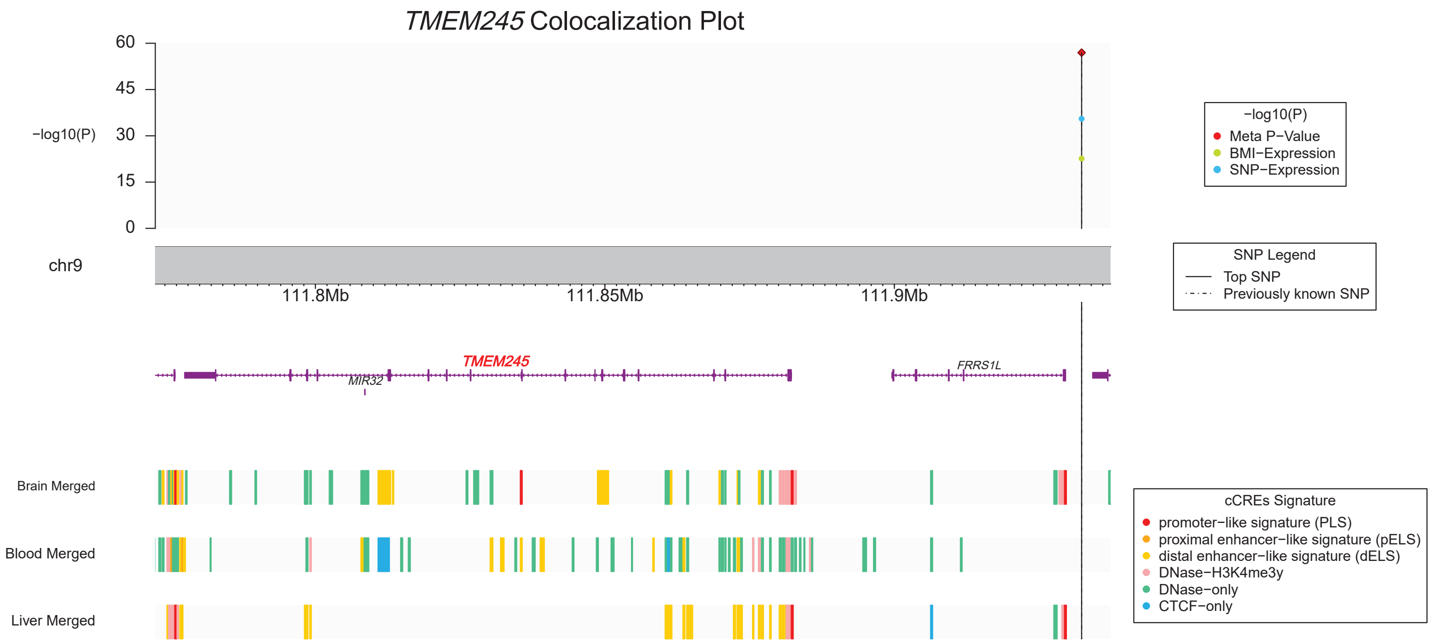

**Supplementary Figure 5.** Regional association plot for the *ZNF646* gene including association results for the discovery sample (Framingham Heart Study) for each tested SNP with gene expression (blue), gene expression with BMI (green), and the correlated meta-analysis for SNP ~ gene expression ~ BMI (red). Annotation for potential ccREs from ENCODE are included for the region.

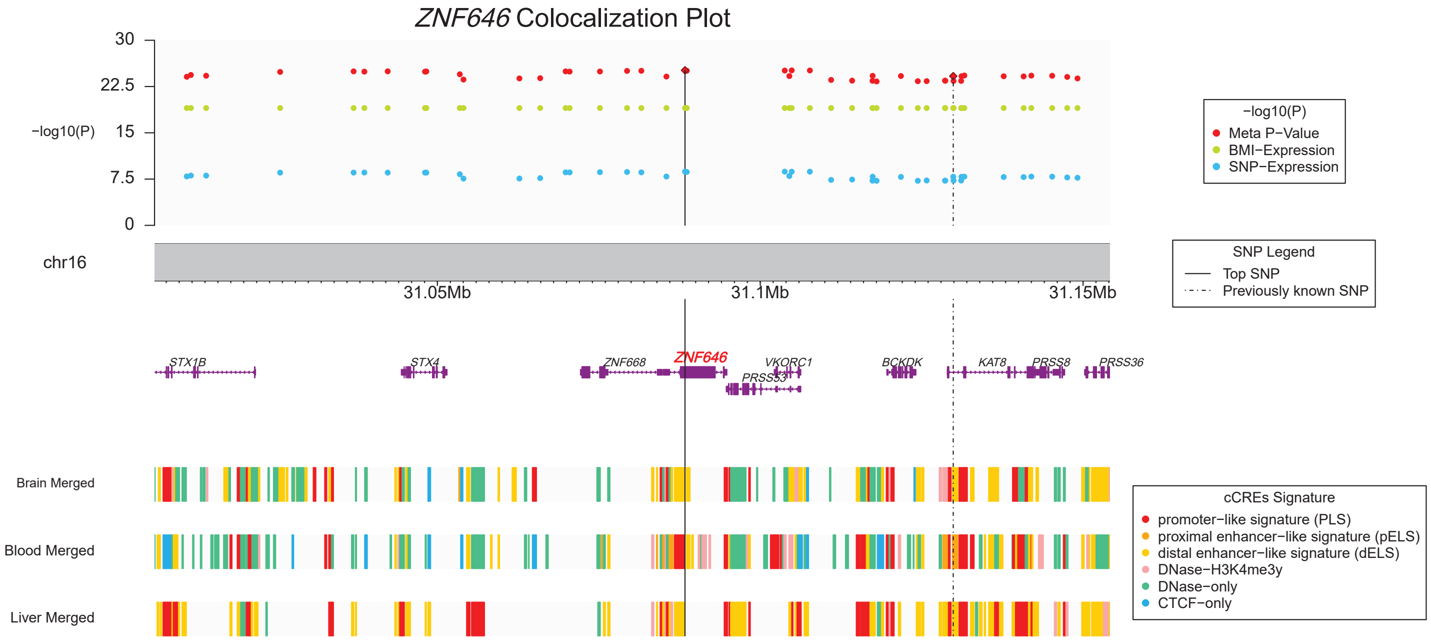
